# Risk factors for adolescent smoking uptake – analysis of prospective data from the UK Millennium Cohort Study

**DOI:** 10.1101/2022.05.26.22275632

**Authors:** Charlotte Vrinten, Jennie C Parnham, Filippos T Filippidis, Nicholas S Hopkinson, Anthony A Laverty

**Affiliations:** Public Health Policy Evaluation Unit, School of Public Health, Imperial College London, England; NHLI, Imperial College, Royal Brompton Hospital Campus, London, UK

**Keywords:** adolescent, cohort studies, peer group, social media, smoking, tobacco use

## Abstract

**Background:** Most smokers take up their habit in adolescence. Risk factors for smoking are changing over time as demographics shift, and technologies such as social media create new avenues for the tobacco industry to recruit smokers. We assessed risk factors associated with smoking uptake among a representative cohort of UK adolescents.

**Methods:** Data come from 8,944 children followed prospectively as part of the UK Millennium Cohort Study. Smoking uptake was assessed as adolescents who had never smoked tobacco at age 14, but reported smoking ≥1 cigarette per week by age 17 (regular smoking). We used logistic regression to assess associations between smoking uptake and selected socio-demographic factors including household income, caregiver smoking, peer smoking, and social media use. Weighted percentages and Office for National Statistics Data were used to estimate numbers of regular smokers and new smokers in the UK.

**Results:** Among the whole sample, 10.6% of adolescents were regular smokers at age 17. Of these, 52% had started smoking between ages 14 and 17. Uptake was more common if caregivers (14.7% vs 5.7%, p<0.001) or friends smoked (13.0% vs. 5.1%, p<0.001), and among those reporting >5hours/day of social media use (10.0% vs 5.2%, p=0.006). Applying these percentages to population data, an estimated 160,000 adolescents in the UK were regular smokers by age 17, of whom more than 100,000 took up the habit between ages 14 and 17.

**Discussion:** Smoking behaviour remains highly transmissible within families and peer groups, reinforcing inequalities. Social media are highlighted as a potential vector.

**What this paper adds:** *What is already known on this subject?:* Cross sectional data suggest that certain population groups, such as those from more deprived backgrounds, are more likely to take up smoking earlier in life, a factor which is linked to greater health risks.

*What important gaps in knowledge exist on this topic?:* There are few prospective studies with nationally representative data that track smoking uptake in adolescence and how this is changing over time in the UK.

*What this study adds:* Around one in nine adolescents in the UK were regular smokers by age 17 and around half (52%) had taken up the habit since they were 14 years old. Smoking among peers and caregivers was linked to adolescent smoking uptake and more time spent on social media was associated with greater likelihood of smoking uptake.

## INTRODUCTION

Most smokers take up the habit during their teenage years and earlier uptake of smoking is linked to being more likely to smoke in later years [1]. Additionally, inequalities between groups in smoking uptake are an important driver of inequalities in tobacco-related health outcomes [2]. The UK Government has committed to achieving a “smoke free generation” and preventing uptake among adolescents will be key to achieving this [3] and the Children’s Charter for Lung Health includes addressing child smoking as one of its key elements [4]. Previous analysis of the UK Millennium Cohort Study (MCS) identified that caregiver and peer smoking were important factors associated with smoking uptake by age 14. As this cohort has now aged, it is possible to investigate factors associated with continued or new uptake of smoking in late adolescence.

In the present paper we therefore extend previous analyses by: 1) assessing levels and risk factors for smoking in late adolescence (17 years), 2) investigating risk factors for smoking uptake between early and late adolescence, and 3) presenting regional estimates of smoking and smoking uptake among late adolescents.

## METHODS

The MCS is a birth cohort study which follows children born between September 2000 and January 2002 [5]. We used data collected at waves six and seven, when the children were approximately 14 and 17 years old, respectively. 9,848 children participated in both waves. After excluding those with missing data, 8,944 (90.8%) individuals were available for analyses.

Smoking status at waves six and seven was assessed by asking children to select one of six statements that best described them: “I have never smoked cigarettes”, “I have only ever tried smoking cigarettes once”, “I used to smoke sometimes but I never smoke a cigarette now”, “I sometimes smoke cigarettes now but I don’t smoke as many as one a week”, “I usually smoke between one and six cigarettes a week”, and “I smoke more than six cigarettes a week”. Regular smoking at age 17 was defined as those who reported smoking at least one cigarette per week at wave seven. Smoking uptake between age 14 and 17 was defined as those who reported “never” smoking at age 14 (wave six) and regular smoking at age 17 (wave seven).

In separate logistic regression models, we assessed associations of age, gender, ethnicity, household income, country, caregiver current smoking, peer smoking, and social media use with regular smoking at age 17 and with smoking uptake between age 14 and 17.

We estimated national and regional numbers of smoking at age 17 and smoking uptake between 14 and 17 years using data on population size by age from the Office for National Statistics (ONS). We used survey weights generated by the survey team to adjust for non-response bias and sampling.

Further details on survey questions and producing national estimates are given in the Appendix.

## RESULTS

About one in nine participants (10.6%) reported regular smoking at age 17 (Table 1). Of these, 52% took up smoking between the ages of 14 and 17, 11% were already smoking regularly at age 14, and 37% had tried smoking or smoked less than one cigarette per week at age 14. Of the never-smokers at age 14, 6.3% (N=488) reported regular smoking (i.e. at least one cigarette per week) at age 17.

**Table 1.** Adjusted logistic regression analyses of regular smoking at age 17 and smoking uptake between age 14 and 17 years (unweighted %; weighted aOR).

|  | Regular smoking at age 17<br>(N=8,944) |  | Smoking uptake between ages 14<br>and 17 (N=7,786)* |  |
| --- | --- | --- | --- | --- |
|  | N (%) | aOR (95% CI)^ | N (%) | aOR (95% CI)^ |
| All | 948 (10.6) | - | 488 (6.3) | - |
| Age |  |  |  |  |
| 16 | 272 (9.3) | Ref | 144 (5.5) | Ref |
| 17/18 | 676 (11.3) | 0.97 (0.78; 1.21) | 344 (6.6) | 0.94 (0.71; 1.25) |
| Gender |  |  |  |  |
| Male | 458 (10.6) | Ref | 256 (6.7) | Ref |
| Female | 490 (10.6) | 0.90 (0.70; 1.16) | 232 (5.9) | 0.81 (0.58; 1.12) |
| Ethnicity |  |  |  |  |
| White | 853 (12.0) | Ref | 448 (7.3) | Ref |
| Mixed | 44 (10.5) | 1.04 (0.57; 1.92) | 13 (3.7) | 0.64 (0.26; 1.58) |
| Indian | 8 (3.1) | 0.35 (0.15; 0.82) | 5 (2.1) | 0.44 (0.16; 1.25) |
| Pakistani/ Bangladeshi | 21 (3.1) | 0.17 (0.10; 0.31) | 12 (1.9) | 0.17 (0.07; 0.38) |
| Black or Black British | 8 (2.8) | 0.27 (0.12; 0.64) | 3 (1.1) | 0.22 (0.06; 0.74) |
| Other | 14 (6.3) | 0.78 (0.38; 1.59) | 7 (3.7) | 0.75 (0.30; 1.91) |
| Household income |  |  |  |  |
| Q1 (highest) | 162 (7.2) | Ref | 89 (4.3) | Ref |
| Q2 | 191 (8.8) | 1.40 (1.01; 1.94) | 103 (5.3) | 1.66 (1.07; 2.57) |
| Q3 | 203 (11.3) | 1.32 (0.97; 1.79) | 123 (7.8) | 1.64 (1.19; 2.26) |
| Q4 | 219 (15.9) | 2.30 (1.73; 3.06) | 96 (8.7) | 2.39 (1.64; 3.47) |
| Q5 (lowest) | 173 (13.0) | 1.79 (1.25; 2.56) | 77 (7.0) | 1.96 (1.16; 3.29) |
| Country |  |  |  |  |
| England | 607 (10.2) | Ref | 295 (5.7) | Ref |
| Wales | 136 (11.4) | 0.84 (0.57; 1.23) | 77 (7.5) | 1.03 (0.64; 1.66) |
| Scotland | 121 (12.8) | 1.22 (0.95; 1.58) | 63 (7.7) | 1.19 (0.84; 1.69) |
| Northern Ireland | 84 (10.1) | 0.97 (0.68; 1.38) | 53 (7.2) | 0.99 (0.68; 1.44) |
| Caregiver smoking |  |  |  |  |
| No | 576 (7.9) | Ref | 324 (5.0) | Ref |
| Yes | 367 (23.2) | 2.06 (1.68; 2.52) | 162 (13.6) | 2.06 (1.57; 2.71) |
| No answer | 5 (7.5) | 1.28 (0.40; 4.13) | 2 (3.3) | 0.76 (0.13; 4.35) |
| Peer smoking |  |  |  |  |
| No | 284 (5.1) | Ref | 229 (4.3) | Ref |
| At least some | 609 (22.9) | 3.67 (2.78; 4.85) | 223 (12.6) | 2.32 (1.62; 3.32) |
| No answer | 55 (7.5) | 1.26 (0.81; 1.94) | 36 (5.4) | 1.01 (0.61; 1.67) |
| Social media use on<br>weekdays |  |  |  |  |
| Less than 1 hour | 189 (6.0) | Ref | 120 (4.1) | Ref |
| Between 1 and 5 hours | 457 (10.9) | 1.38 (1.05; 1.81) | 253 (6.9) | 1.41 (1.01; 1.96) |
| More than 5 hours | 302 (18.8) | 1.91 (1.41; 2.59) | 115 (9.8) | 1.69 (1.16; 2.46) |

\*This is a subsample of the entire sample and consists of “never smokers” at age 14 (i.e. it excludes those who reported any smoking at age 14).
^aOR= adjusted odds ratio, mutually adjusted for all covariates included in the table. 95% CI = 95% confidence interval.

Factors associated with being a regular smoker at age 17 and taking up smoking between ages 14 and 17 were similar (Table 1). Those from ethnic minority backgrounds were less likely to be regular smokers at age 17 or to take up smoking, while those from lower income households, and those with caregivers and peers who smoked were more likely to do so.

Adolescents whose caregiver was smoking when they were 14 were more than twice as likely to be a smoker aged 17, and to start smoking between 14 and 17, than those whose caregivers were not smoking. Similarly, adolescents who reported peer group smoking were more than three times as likely to smoke at age 17 and more than twice as likely to take up smoking between ages 14 and 17 than those whose peers did not smoke. Both regular smoking and smoking uptake were more common among adolescents in lower income households. For example, those in the lowest household income group were almost twice as likely to take up smoking as those in the highest household income group (AOR = 1.96, 95% CI 1.16 to 3.29).

Those who spend 1-5 hours per day on social media were 1.4 times more likely, and those who spend more than 5 hours were almost twice as likely, to smoke at age 17 or take up smoking between 14 and 17 than those who spent less than 1 hour per day on social media (test for trend p<0.001 for both outcomes)(Table 1).

Our weighted estimates show that more than 160,000 adolescents in the UK were regular smokers by age 17, of whom more than 100,000 took up the habit between ages 14 and 17 (Appendix Table 2). Between the countries of the UK, smoking uptake ranged from 7.0% in England 8.6% in Wales.

## Discussion

Data from the Millennium Cohort Study show that of the nearly one in nine adolescents in the UK who were regular smokers by age 17, around half (52%) had taken up the habit since they were 14 years old. Caregiver smoking, peer smoking, and social media use were linked to uptake of tobacco smoking among UK adolescents.

Previous analyses of the same cohort at age 14 found that 1.9%, or an estimated 39,653 early teens around the UK were smokers [6]. Together, these findings indicate that a large group of UK adolescents still take up smoking despite the government’s pledge to create a “smoke-free generation” and that approaches to address this need to be delivered across childhood. They also serve as a reminder of the transmissibility of the smoking epidemic with peer and caregiver smoking increasing tobacco use among adolescents. Our findings that adolescents in lower income households were more likely to take up smoking and to be regular smokers highlight the inequalities in smoking harms and the need for a systems approach in tackling tobacco use [7] [8].

We found a significant independent association between social media use and smoking uptake. This finding is in line with other research, mainly conducted in the USA, which has found, for example, increased susceptibility to smoking uptake and higher levels of smokeless tobacco use among children exposed to online tobacco advertising [9][10]. Although causation cannot be determined from these findings, they heighten existing concerns that social media content may promote smoking, reinforcing the call from the Royal College of Physicians to ban all social media marketing of tobacco products [11][12]. These findings also strengthen arguments that legislation to address online safety should consider public health harms, including those from tobacco advertising, and of the need for continued awareness over the changing landscape of tobacco advertising over time [13].

A novelty of this study is that our covariates, such as household income, caregiver smoking, peer smoking, and social media use, were assessed prospectively, before uptake of smoking, adding strength to the temporality of the relationship between these factors and subsequent smoking uptake. Limitations to this work include that smoking measures were based on self-report, but previous studies have shown that this is a reliable indicator for the prevalence of actual smoking behaviour [14]. Furthermore, we did not consider e-cigarette use in the analyses, although an estimated 5-8% of adolescents use these [7]. Hence, we may have underestimated total use of nicotine-containing products by adolescents. Our findings regarding social media use are limited by the fact that the measure used was hours of use, and not a more specific measure such as actual exposure to pro-tobacco advertisements or messages. Finally, while cohort studies are prone to attrition over time, we used the survey weights provided to adjust for this and to ensure population-representativeness.

In conclusion, these prospective data show that the relationship of caregiver and peer group smoking with smoking uptake persists throughout childhood, and highlights a potential role for social media as an important novel vector.

## Supporting information

Research checklist (STROBE)

## Data Availability

All data produced are available online at https://ukdataservice.ac.uk

