## Appendix for "Risk factors for adolescent smoking uptake – analysis of prospective data from the UK Millennium Cohort Study"

**Appendix contents**

1. Additional description of the Millennium Cohort Study and sample selection
2. Assessment of outcomes
3. Assessment of covariates
4. Population data
5. Analyses
6. Additional analyses
7. References
8. Appendix tables

**1. Additional description of the Millennium Cohort Study and sample selection**

The original sample of the Millennium Cohort Study (MCS) consisted of nearly 19,000 children who were born in England, Wales, Scotland and Northern Ireland between September 2000 and January 2002. A stratified, clustered random sample design was used with oversampling from disadvantaged areas (i.e. those living in the poorest 25% of wards (or Electoral Divisions in Wales) according to the Child Poverty Index) and areas with a high proportion of ethnic minority groups (at least 30%). Once the sample wards were selected, all children on the Child Benefit register who turned 9 months olds during the survey period were invited to participate.

Seven waves of data collection have taken place since inception of the cohort, at age 9 months and 3, 5, 7, 11, 14 and 17 years. Data for wave six (14 years) were collected between Jan 2015 and March 2016. Data for wave seven (17 years) were collected between Jan 2018 and March 2019 (Fitzsimons et al., 2020).

Wave six resulted in 11,726 families with productive interviews and 10,625 for wave seven. Almost ten thousand children completed both waves 6 and 7 (N=9,848). Of these, 901 (9.1%) had missing data on smoking at wave six and/or wave seven and three participants (0.03%) had missing data on social media use at wave six, and these were excluded from our analytical sample. This resulted in an analytical sample of N=8,944.

Descriptive statistics for the sample can be found in Appendix Table 1.

**2. Assessment of outcomes**

Smoking status was assessed by asking children to select one of six statements that best described them at waves six and seven: "I have never smoked cigarettes", "I have only ever tried smoking cigarettes once", "I used to smoke sometimes but I never smoke a cigarette now", "I sometimes smoke cigarettes now but I don't smoke as many as one a week", "I usually smoke between one and six cigarettes a week", and "I smoke more than six cigarettes a week".

*Regular smoking at age 17* was defined as those who reported smoking at least one cigarette per week at wave seven. *Smoking uptake between age 14 and 17* was defined as those who reported "never" smoking at age 14 (wave six) and regular smoking at age 17 (wave seven).

**3. Assessment of covariates**

Age at wave 7, gender (wave 7), ethnicity in six groups (wave six), and UK country of residence (wave seven) were recorded in the survey. For missing data on age at wave 7 (n=107, 1.2%), gender (n=107, 1.2%), ethnicity (n=62, 0.7%), and region of residence (n=107, 1.2%), we carried forward responses from previous survey waves (for age at wave 7, this was imputed as age at wave 6 plus 3 years). For the logistic regression analyses, we collapsed the categories for age 17 and 18 at wave seven due to low numbers of participants aged 18 (n=28; 0.3%). Missing data on parental (n=62, 0.8%) and peer smoking (n=730, 8.2%) were coded as separate category for analysis.

Household income (wave six) was based on the Organisation for Economic Co-operation and Development (OECD) equivalised income and was categorised in five groups (Fitzsimons, 2020).

Caregiver smoking (yes/no) at wave six was assessed in the interview with the cohort member's main carer.

Peer smoking at wave six was assessed in the cohort member interview with the question: "How many of your friends smoke cigarettes? Do not include e-cigarettes" with response options: "none of them", "some of them", "most of them", "all of them". These were recoded for analysis as "No" ("none of them") versus "At least some" (all other responses). Missings were recoded as a separate category for analysis.

Social media use at wave six was assessed with the question: "On a normal week day during term time, how many hours do you spend on social networking or messaging sites or Apps on the internet such as Facebook, Twitter and WhatsApp?" with response options: "none", "less than half an hour", "half an hour to less than 1 hour", "1 hour to less than 2 hours", "2 hours to less than 3 hours", "3 hours to less than 5 hours", "5 hours to less than 7 hours", "7 hours or more". Responses were recoded for analysis as "less than 1 hour", "between 1 and 5 hours", and "more than 5 hours".

**4. Population data**

We used mid-2018 estimates from the Office for National Statistics (ONS) on population size by single year of age and sex (Office for National Statistics, 2019). This was matched to the MCS data based on UK country of residence and also on Government Office Region for England.

We used ONS data on population size for 16- and 17-year olds by country and region, and weighted percentages for regular smokers and smoking uptake among 16- and 17-year olds by country/region obtained from the MCS data to produce regional estimates for the absolute number of adolescents (with 95% confidence intervals) who were regular smokers by age 17 and for the absolute number of adolescents (with 95% CI) who had taken up smoking between age 14 and 17.

Due to small numbers in the MCS data and consequent potential for unreliable estimates, we excluded 18 year olds from these analyses (n=28, 0.3%).

### 5. Analyses

We used unweighted data for the descriptive characteristics of the samples, and used the weights provided by the Millennium Cohort Study team to adjust for non-response bias (from study attrition) and sampling for the logistic regression analyses of smoking at age 17, smoking uptake between age 14 and 17, and to calculate the country/regional estimates of smokers at age 17 and smoking uptake between age 14 and 17. More details on the construction of these weights can be found in the Millennium Cohort Study Seventh Sweep (MCS7): Technical Report (Centre for Longitudinal Studies, 2019) and the Millennium Cohort Study Age 17 Sweep (MCS7): User Guide (Fitzsimons et al., 2020).

### 6. Additional analyses

Appendix Table 2 shows the estimates of adolescent smoking at age 17 and smoking uptake between age 14 and 17 (percentages and absolute numbers), for the four constituent countries of the UK and government office regions of England.

Appendix Figure 1. Presents a graphical representation of the adjusted odds ratios and 95% confidence intervals of regular smoking at age 17 and smoking uptake between age 14 and 17 as reported in the main manuscript.

**8. Appendix tables.**

Appendix Table 1. Descriptive sample characteristics (unweighted) of the whole sample (N=8,944), and the subsamples of those who had never smoked at age 14 (N=7,786) and those who had ever smoked at age 14 (N=1,158).

| | All (N=8,944) | Never smokers age 14 (N=7,786) | Ever smokers age 14 (N=1,158) |
|---|---|---|---|
| | **N (%)** | **N (%)** | **N (%)** |
| Age at wave 7 | | | |
| 16 | 2,933 (32.8) | 2,604 (33.4) | 329 (28.4) |
| 17/18 | 6,011 (67.2) | 5,182 (66.6) | 829 (71.6) |
| Gender | | | |
| Male | 4,331 (48.4) | 3,829 (49.2) | 502 (43.4) |
| Female | 4,613 (51.6) | 3,957 (50.8) | 656 (56.7) |
| Ethnicity | | | |
| White | 7,087 (79.2) | 6,117 (78.6) | 970 (83.8) |
| Mixed | 420 (4.7) | 352 (4.5) | 68 (5.9) |
| Indian | 257 (2.9) | 243 (3.1) | 14 (1.2) |
| Pakistani and Bangladeshi | 669 (7.5) | 617 (7.9) | 52 (4.5) |
| Black or Black British | 290 (3.2) | 265 (3.4) | 25 (2.2) |
| Other | 221 (2.5) | 192 (2.5) | 29 (2.5) |
| Household income | | | |
| Q1 (highest) | 2,263 (25.3) | 2,072 (26.6) | 191 (16.5) |
| Q2 | 2,178 (24.4) | 1,936 (24.9) | 242 (20.9) |
| Q3 | 1,801(20.1) | 1,569 (20.2) | 232 (20.0) |
| Q4 | 1,375 (15.4) | 1,105 (14.2) | 270 (23.3) |
| Q5 (lowest) | 1,327 (14.8) | 1,104 (14.2) | 223 (19.3) |
| Country | | | |
| England | 5,970 (66.8) | 5,199 (66.8) | 771 (66.6) |
| Wales | 1,191 (13.3) | 1,028 (13.2) | 163 (14.1) |
| Scotland | 949 (10.6) | 819 (10.5) | 130 (11.2) |
| Northern Ireland | 834 (9.3) | 740 (9.5) | 94 (8.1) |
| Parental smoking | | | |
| No | 7,292 (81.5) | 6,538 (84.0) | 754 (65.1) |
| Yes | 1,585 (17.7) | 1,188 (15.3) | 397 (34.3) |
| No answer | 67 (0.8) | 60 (0.8) | 7 (0.6) |
| Peer smoking | | | |
| No | 5,554 (62.1) | 5,350 (68.7) | 204 (17.6) |
| At least some | 2,660 (29.7) | 1,764 (22.7) | 896 (77.4) |
| No answer | 730 (8.2) | 672 (8.6) | 58 (5.0) |
| Social media use on weekdays | | | |
| Less than 1 hour | 3,150 (35.2) | 2,947 (37.9) | 203 (17.5) |
| Between 1 and 5 hours | 4,183 (46.8) | 3,663 (47.1) | 520 (44.9) |
| More than 5 hours | 1,611 (18.0) | 1,176 (15.1) | 435 (37.6) |
| Smoking at age 14 | | | |
| Never | 7,786 (87.0) | 7,786 (100.0) | --- |
| Ever smoking but not weekly | 1,017 (11.4) | --- | 1,017 (87.8) |
| Regular (at least weekly) | 141 (1.6) | --- | 141 (12.2) |
| Regular smoking at age 17 | | | |
| No | 7,996 (89.4) | 7,298 (93.7) | 698 (60.3) |
| Yes | 948 (10.6) | 488 (6.3) | 460 (39.7) |

Appendix Table 2. Regional estimates for regular smoking at age 17 and smoking uptake between age 14 and 17 (using weighted data).

| | Adolescents who are regular smokers at age 17 | | Adolescents taking up smoking between age 14 and 17 * | |
|---|---|---|---|---|
| Country/Region | % (95% CI)^ | N (95% CI)^ | % (95% CI)^ | N (95% CI)^ |
| England | 11.4 (10.0; 12.9) | 137961 (120839; 155082) | 7.0 (5.7; 8.2) | 83878 (68452; 99303) |
| North East | 11.3 (4.8; 17.8) | 6186 (2625; 9748) | 6.6 (2.3; 10.9) | 3597 (1260; 5934) |
| North West | 11.7 (7.6; 15.8) | 18546 (12054; 25037) | 7.9 (5.2; 10.6) | 12518 (8272; 16764) |
| Yorkshire and the Humber | 9.5 (6.9; 12.0) | 11281 (8262; 14299) | 4.9 (3.2; 6.6) | 5866 (3849; 7883) |
| East Midlands | 8.9 (5.8; 11.9) | 9085(5930; 12241) | 3.6 (1.6; 5.6) | 3712 (1655; 5768) |
| West Midlands | 10.9 (7.8; 14.1) | 14440 (10332; 18548) | 7.4 (4.0; 10.7) | 9712 (5281; 14143) |
| East of England | 11.2 (5.9; 16.5) | 15146 (7990; 22301) | 7.8 (2.3; 13.3) | 10541 (3143; 17939) |
| London | 8.4 (5.1; 11.7) | 15672 (9510; 21833) | 5.1 (2.2; 8.0) | 9490 (4086; 14894) |
| South East | 14.3 (11.4; 17.2) | 28990 (23123; 34856) | 9.6 (6.9; 12.4) | 19490 (13874; 25106) |
| South West | 14.1 (9.4; 19.0) | 16367 (10834; 21901) | 7.1 (3.5; 10.7) | 8193 (4070; 12316) |
| Wales | 11.0 (8.4; 13.5) | 7359 (5643; 9075) | 8.6 (5.9; 11.3) | 5760 (3924; 7595) |
| Scotland | 13.0 (10.6; 15.4) | 14213 (11572; 16854) | 8.0 (6.0; 10.0) | 8740 (6540; 10940) |
| Northern Ireland | 10.8 (8.5; 13.2) | 4863 (3798; 5928) | 7.2 (5.1; 9.2) | 3219 (2302; 4136) |
| Overall | 11.5 (10.3; 12.7) | 164313 (146815; 181811) | 7.1 (6.0; 8.2) | 101715 (85994; 117435) |

*These estimates are for "never smokers" at age 14 who reported regular smoking at age 17.

^ 95% CI = 95% confidence interval

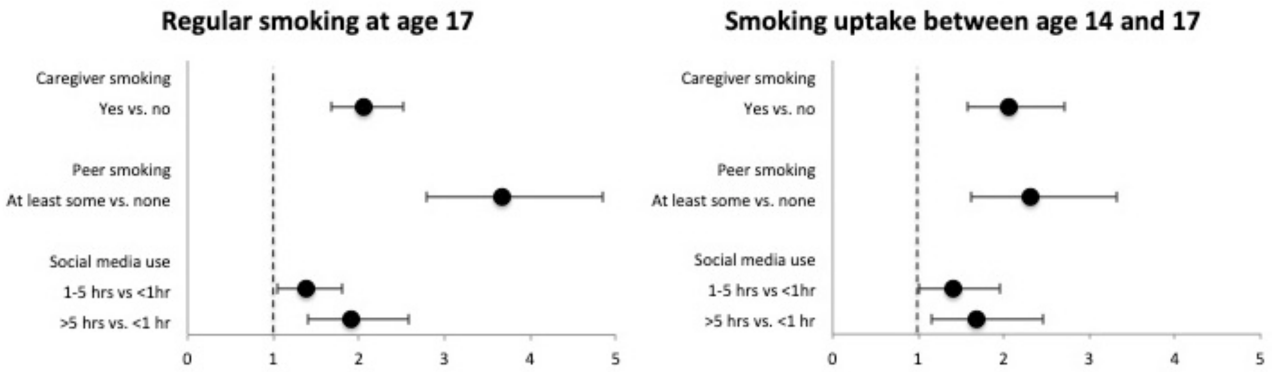


Appendix Figure 1. Adjusted odds ratios and 95% confidence intervals of regular smoking at age 17 and smoking uptake between age 14 and 17.
